# Prevalence of new onset diabetes mellitus among patients with pancreatic ductal adenocarcinoma: a population-based study

**DOI:** 10.64898/2026.08.05.26359758

**Authors:** Jan Bures, Katerina Hejcmanova, Tereza Dianova, Ondrej Ngo, Darina Kohoutova, Radek Pohnan, Jan Skrha, Stepan Suchanek, Petr Urbanek, Ladislav Dusek, Miroslav Zavoral, Ondrej Majek

**Affiliations:** Institute of Gastrointestinal Oncology, Military University Hospital Prague, Czech Republic; Department of Medicine, Charles University, First Faculty of Medicine, Prague and Military University Hospital Prague, Czech Republic; Biomedical Research Centre, University Hospital Hradec Kralove, Czech Republic; National Screening Centre, Institute of Health Information and Statistics of the Czech Republic, Prague, Czech Republic; Institute of Biostatistics and Analyses, Masaryk University, Faculty of Medicine, Brno, Czech Republic; The Royal Marsden NHS Foundation Trust, London, United Kingdom; Department of Surgery, Charles University, Second Faculty of Medicine, Prague and Military University Hospital Prague, Czech Republic; Third Department of Internal Medicine – Endocrinology and Metabolism, Charles University, First Faculty of Medicine, Prague and General University Hospital in Prague, Czech Republic; RECETOX, Faculty of Science, Masaryk University, Brno, Czech Republic

**Keywords:** new-onset diabetes mellitus, prediabetes, pancreatic cancer, prevalence

## Abstract

**Background:** Pancreatic ductal adenocarcinoma (PDAC) has remained one of the most serious malignancies and is still a leading cause of cancer-related deaths worldwide. Great attention has been paid to the relationship between diabetes mellitus and PDAC. The aim of our study was to analyse the mutual association of PDAC and diabetes mellitus in the entire population of the Czech Republic within a 5-year period.

**Methods:** The incidence of PDAC in 2018-2022 was estimated based on the individual population data from the Czech National Cancer Registry. Another data source, the National Registry of Reimbursed Health Services that collects data from health insurance companies, was used to identify individuals recently diagnosed with diabetes mellitus. For the purpose of this study, diagnosis of new-onset of diabetes mellitus was defined as the time of the first prescription of any antidiabetic drug or another related health care service. Subsequently, patients diagnosed with PDAC in 2022 were followed retrospectively to see if they had been diagnosed with diabetes mellitus in the last five years before diagnosis of pancreatic cancer.

**Results:** In 2022, 2,189 patients aged 60 years or older were diagnosed with pancreatic cancer. New-onset diabetes was observed in 17.4% within five years prior to diagnosis, with the highest occurrence (12.4%) within the last three years. Among patients aged 60-74 years, the respective proportions were 14.4% within three years and 5.7% four to five years prior to pancreatic cancer diagnosis.

**Conclusion:** The incidence of pancreatic cancer in the Czech Republic is among the highest in Europe. One-fifth of PDAC cases are diagnosed following new-onset diabetes mellitus in patients over sixty. Unintended significant weight loss combined with new-onset diabetes thus must not be overlooked, as these can be early signs of PDAC. An individualised diagnostic work-up should follow without any unnecessary delay.

**Note:** The study was presented in part, as an abstract and moderated poster, at the 32nd United European Gastroenterology Week, Vienna, October 12-15, 2024 [abstract: UEG Journal, 2024(12), S8: 565-566].

**Key Summary:**

- Summarise the established knowledge on this subject
  - Pancreatic ductal adenocarcinoma (PDAC) is□one of the most lethal□gastrointestinal cancers, with an increasing incidence worldwide.
  - The mutual association between□PDAC and diabetes mellitus has been recognized for decades. While diabetes mellitus can be a risk factor for pancreatic cancer, new-onset diabetes□often serves as□an early sign of PDAC.
  - Different types of diabetes mellitus□are□associated with PDAC. Type 2 diabetes is the most common (characterized by increased insulin secretion and receptor resistance). Secondary diabetes mellitus in non-malignant pancreatic diseases (e.g., chronic pancreatitis) involves a reduction of islet-cell mass, leading to decreased insulin secretion□but□normal insulin sensitivity.□Type 3c diabetes associated with PDAC is considered□a paraneoplastic syndrome, characterized by decreased insulin secretion and increased insulin resistance.□
- What are the significant and/or new findings of this study?
  - Our study was based on data from Czech national healthcare registries (2018– 2022).
  - The age-standardized incidence rate of PDAC in the Czech Republic was 24.6 per 100,000 population in 2023.
  - In 2022, 2,189 patients over the age of 60 were diagnosed with PDAC. New-onset diabetes mellitus was recorded in 12.4% of patients within three years□prior to their□cancer diagnosis.
  - Specifically, among□patients aged 60–74 years, this proportion was 14.4% (three years prior to diagnosis),□reaching a total of□20.1% over a five-year period.
  - Unintended significant weight loss combined with new-onset diabetes must not be overlooked, as these can be early signs of PDAC.
  - An individualized diagnostic work-up should follow without□unnecessary delay

## INTRODUCTION

Pancreatic ductal adenocarcinoma (PDAC) belongs among the most fatal gastrointestinal cancers, with an increasing incidence worldwide [1,2]. The global burden of pancreatic cancer has doubled in the past 25 years [3]. Despite all the enormous effort in research, diagnostics and treatment, the prognosis of PDAC remains very poor (5-year survival rate only about 5-10%) [1,4].

The relationship between pancreatic cancer and diabetes mellitus has been acknowledged for centuries. Italian anatomist Giovanni Battista Morgagni was given credit for his early observations of mutual association of pancreatic cancer and diabetes mellitus in 1761 [5]. Richard Bright (Guy’s Hospital London) in 1833 described a patient with newly diagnosed diabetes mellitus who died of pancreatic cancer six months later [6]. The first Czech case reports on diabetes in pancreatic cancer came in the mid 1930s [7]. Czech internist Jiri Syllaba described in 1934 a case of new-onset diabetes mellitus preceding obstructive jaundice in pancreatic cancer by a few months [8]. Systematic attention to the relationship between pancreatic cancer and diabetes mellitus has been paid since the 1950s [9]. The first meta-analysis between pancreatic cancer and diabetes mellitus was conducted in 1995. It analysed 20 appropriate out of 30 available case-control and cohort studies published worldwide and reported an overall estimated relative risk of pancreatic cancer associated with diabetes mellitus of 2.1 [10].

Different types of diabetes mellitus can be associated with PDAC. Type 2 diabetes is the most common (with increased secretion of insulin and increased receptor resistance to insulin). Secondary diabetes mellitus in non-malignant diseases of the pancreas (e.g. chronic pancreatitis), with a reduction or loss of islet-cell mass, has decreased secretion of insulin and normal sensitivity to insulin. Type 3c is a paraneoplastic syndrome with decreased secretion of insulin and increased resistance to insulin [see ref. 11 for details].

At the time of diagnosis of pancreatic cancer, impaired glucose metabolism has been reported to be present in variable rates according to particular studies [11]. The aim of our study was to analyse the mutual relationship of pancreatic cancer and diabetes mellitus in the entire population of the Czech Republic throughout a 5-year period.

## MATERIAL AND METHODS

### Data sources

The analysis was based on patient-level data from the National Health Information System, enabling secondary use of official Czech national healthcare data [12, 13]. Data on the incidence of pancreatic cancer in recent years was provided by the Czech National Cancer Registry. Another data source, the National Registry of Reimbursed Health Services, which collects data on reimbursed care from health insurance companies in the Czech Republic, was used to identify people recently diagnosed with diabetes mellitus. For the purpose of this study, diagnosis of new-onset of diabetes mellitus was defined as the time of the first prescription of any antidiabetic drug (the subgroup of patients with prediabetes treated with metformin was included), and/or the first medical procedures connected with diabetes, and/or the first reported health conditions associated with diabetes mellitus. The detailed list of procedure and diagnosis codes used to define diabetes mellitus is provided in Supplementary Table S1. A sensitivity analysis was additionally performed using an alternative definition of diabetes mellitus based exclusively on antidiabetic drug prescriptions. The results of this analysis are shown in Supplementary Table S2.

### Statistical methods and analyses

Before conducting the basic analysis, the epidemiological situation regarding diabetes mellitus and pancreatic cancer in the Czech Republic was monitored.

The analyses focus on a retrospective view of individuals diagnosed with pancreatic cancer in 2022. These individuals were monitored so that it could be determined whether they had also been diagnosed with diabetes mellitus within a defined time interval prior to this diagnosis. The age category of individuals was related to their age at the time of diagnosis of pancreatic cancer.

### Ethics

The analysis was conducted using data from national health registries whose establishment and operation are governed by a legal framework designed to regulate the collection, processing and sharing of health information. This legal framework includes the following legislation:

- Act No. 372/2011 Coll., on Health Services and Conditions of Their Provision,
- Act No. 325/2021 Coll., on the Digitization of Health Care,
- Act No. 89/1995 Coll., on the National Statistical Service,
- Act No. 285/2002 Coll., on the Donation, Procurement and Transplantation of Tissues and Organs, and on Amendments to Certain Acts,
- Act No. 258/2000 Coll., on the Protection of Public Health,
- Regulation (EU) 2016/679, General Data Protection Regulation (GDPR) and Amendments to Related Acts.

All data handling activities are carried out under the supervision of the Data Handling Committees of the Institute of Health Information and Statistics, which include a designated Data Protection Officer responsible for oversight of work with sensitive data.

## RESULTS

Between 2020 and 2024, a total of 349,570 individuals were newly identified with diabetes mellitus (DM) based on antidiabetic medication records (ATC group A10), of whom 195,023 (55.8%) were aged 60 years or older. Using the extended definition, which included diagnosis codes, healthcare procedures, and diabetes-related medical devices in addition to medication records, 596,956 individuals were identified during the period 2020-2024 (see Figure 1). As pharmacological treatment was not required for classification, this group may include individuals managed with diet alone as well as those with prediabetes.

**Figure 1:**
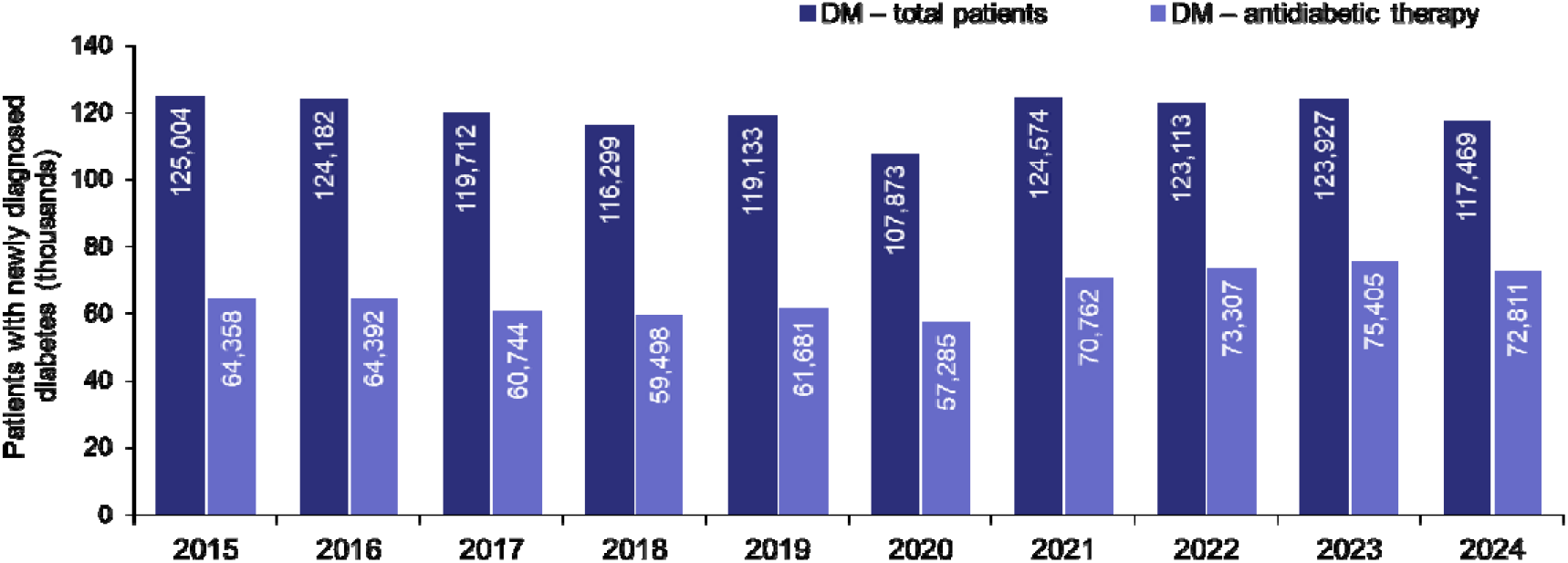
Annual incidence of diabetes mellitus in the Czech Republic, 2015-2024 Data source: National Registry of Reimbursed Health Services Note: Diabetes mellitus (DM) was identified using diagnosis codes (ICD-10), antidiabetic medications (ATC A10), healthcare procedure codes, and diabetes-related medical devices. Presence of at least one criterion was sufficient for classification. “DM – total patients” includes all patients meeting any of these criteria, while “DM – antidiabetic therapy” includes only patients with at least one record of treatment with insulin and/or oral antidiabetic medications (ATC groups A10A and/or A10B). Patients without any recorded antidiabetic treatment may be managed with diet alone and/or may have prediabetes.

Between 2019 and 2023, a total of 12,960 individuals were newly diagnosed with pancreatic cancer in the Czech Republic, corresponding to an average of 2,592 cases per year. The age-standardized incidence rate in 2023 was 24.6 per 100,000 population (see Figure 2).

**Figure 2:**
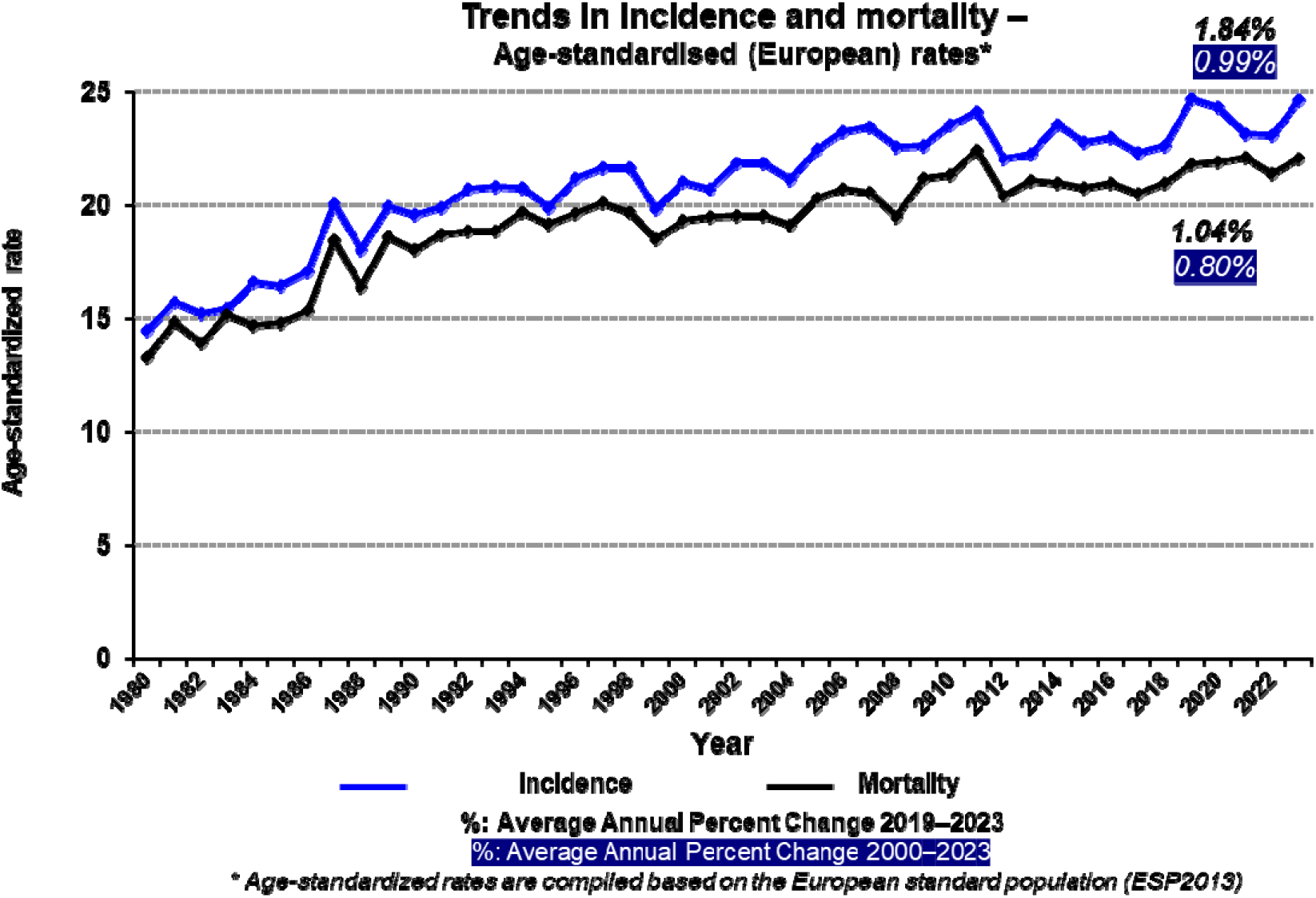
Epidemiology of pancreatic cancer in the Czech Republic, 1980-2023 Data source: Czech National Cancer Registry, Czech Statistical Office

In 2022, 2,189 patients over the age of 60 years were diagnosed with pancreatic cancer. New-onset diabetes mellitus was recorded in 12.4% of patients within three years prior to the cancer diagnosis, and in an additional 5.0% of patients four to five years before diagnosis, corresponding to 17.4% within five years overall. When focusing specifically on patients aged 60-74 years, these proportions were 14.4% and 5.7%, respectively, summing to 20.1% over the five-year period. The mutual association between diabetes mellitus and pancreatic cancer is presented in Table 1. Corresponding results using an alternative definition of diabetes mellitus based on antidiabetic medications is provided in Supplementary Table S2.

**Table 1.** Recent diabetes mellitus in people diagnosed with pancreatic cancer in 2022.

| **Time of diagnosis of diabetes mellitus (DM)** | **39 years and under** | **40-59 years old** | **60-74 years old** | **75 years and over** | **TOTAL** |
| --- | --- | --- | --- | --- | --- |
| DM diagnosed within 1 year prior to cancer diagnosis | 2 (9.6%) | 28 (8.3%) | 98 (8.4%) | 56 (5.5%) | 184 (7.2%) |
| DM diagnosed within 2 years prior to cancer diagnosis | 0 (0.0%) | 9 (2.7%) | 38 (3.3%) | 22 (2.2%) | 69 (2.7%) |
| DM diagnosed within 3 years prior to cancer diagnosis | 0 (0.0%) | 8 (2.4%) | 32 (2.7%) | 26 (2.5%) | 66 (2.6%) |
| DM diagnosed within 4 years prior to cancer diagnosis | 1 (4.8%) | 10 (3.0%) | 37 (3.2%) | 25 (2.4%) | 73 (2.9%) |
| DM diagnosed within 5 years prior to cancer diagnosis | 0 (0.0%) | 7 (2.1%) | 30 (2.6%) | 17 (1.7%) | 54 (2.1%) |
| DM diagnosed 6 or more years prior to cancer diagnosis | 1 (4.8%) | 54 (16.0%) | 417 (35.7%) | 437 (42.8%) | 909 (35.7%) |
| DM diagnosed after the cancer diagnosis | 1 (4.8%) | 15 (4.5%) | 38 (3.3%) | 24 (2.3%) | 78 (3.1%) |
| DM not diagnosed | 16 (76.2%) | 206 (61.1%) | 477 (40.9%) | 415 (40.6%) | 1,114 (43.7%) |
| **TOTAL NUMBER OF PATIENTS DIAGNOSED WITH PANCREATIC CANCER (2022)** | **21 (100.0%)** | **337 (100.0%)** | **1,167 (100.0%)** | **1,022 (100.0%)** | **2,547 (100.0%)** |

## DISCUSSION

Our study focusing on the mutual relationship of new-onset diabetes mellitus and pancreatic cancer was based on the Czech national health-care registries.

Electronic health records offer opportunities to use longitudinal and cumulative robust data for detailed population analyses [14]. And indeed, our current study brought important novel data on this relationship. All subjects with a prescription of any antidiabetic drug were analysed so that patients with prediabetes treated with metformin were enrolled, too. The analysis also included individuals for whom the diagnosis of diabetes mellitus was made based on related reported care or medical devices. Some of these individuals are likely to be in the prediabetes stage. We analysed not only the 3-year time-span of new-onset diabetes prior to cancer diagnosis but also the intervals of 4 and 5 years. New-onset diabetes mellitus in the interval 4-5 years prior to the pancreatic cancer diagnosis was revealed in 5% of individuals over 60 years. We can hypothesize that this was owing to preventive examinations of the Czech general population every other year, which have been long-established in the regulations of the Ministry of Health – the preventative examinations include fasting glycaemia, so that asymptomatic diabetes or prediabetes are recognized earlier. Our findings led to consideration of the introduction of an early detection programme for pancreatic cancer in the Czech Republic based on new-onset of diabetes mellitus in subjects over 60, coupled with significant unintended weight loss (more than 10% body weight within the last 12 months) and other alarm symptoms and/or other possible risk factors.

The National Cancer Institute has published easy-to-understand retrospective and prospective figures which are similar to ours: about 1 in 100 individuals with new onset diabetes mellitus is diagnosed with cancer within three years of their diabetes diagnosis. One in four people diagnosed with pancreatic cancer were first diagnosed with diabetes [15]. Several studies supported this statement.

Olmsted County Study investigated 2,122 subjects over 50 years of age with new-onset diabetes mellitus [16]. PDAC was revealed in 18 patients (0.85%) within 3 years after diabetes was diagnosed (10/18 in the first 6 months). The pancreatic cancer was surgically resectable in one case only. New-onset diabetes was associated with an 8-times higher risk of PDAC compared to the general population of the same age and adjusted to gender (Iowa SEER data) [16]. The same ratio (0.8%) was found in our current large general population-based study, too.

A Nurses’ Health Study and a Health Professionals Follow-Up Study (with repeated assessments over 30 years) enrolled 112,818 women and 46,207 men. In total, 1,116 incident cases of pancreatic cancers were identified [17]. New-onset diabetes mellitus (HR 2.97) accompanied by weight loss (>4 kg; HR 6.75) was associated with a substantially increased risk of developing pancreatic cancer. Older age, previous “healthy weight” and no intentional weight loss elevated this risk further [17,18].

In general, according to several other studies [reviewed by ref. 11], new-onset diabetes (< 2-year duration), older age, higher body-mass index (before the illness) followed by an unintentional weight loss, and a greater frequency of family history of diabetes mellitus were further discriminating features [11,19,20].

However, not all available epidemiological data fully correspond. A large Japanese project (212,775 diabetics; 1,755 pancreatic cancers; period 2012-2021) found no significant association of diabetes mellitus and the onset of pancreatic cancer [21].

Czech Republic has the highest incidence of pancreatic cancer in Europe, together with Hungary and Slovakia [1]. A large Hungarian 10-year population study [22]which included nearly 3,700,000 individuals with diabetes mellitus observed similar results to ours. Diabetes mellitus was associated with a significantly higher risk of pancreatic cancer (HR 2.29). The study’s authors have suggested the inclusion of new-onset diabetes mellitus into a diagnostic algorithm of different cancers [22].

Several new directions had been suggesting that an END-PAC approach is a very important one [23,24]. It is based on the use of four simple values: fasting glycaemia one year before onset of diabetes, fasting glycaemia at the onset of diabetes, body weight change over one year before onset of diabetes, and age at the onset of diabetes mellitus [23]. This score assists in stratifying the risk of PDAC in subjects over 50 with new-onset diabetes mellitus (low-risk: < 0 points; high-risk: ≥ 3 points). A subsequent diagnostic algorithm was proposed [24]. END-PAC was already validated in the USA [25,26] as well as in the UK to predict PDAC [27,28]. Its feasibility has been evaluated in primary care in UK [29]. The updated ENDPAC model uses changes in glycated haemoglobin (HbA1c) concentrations instead of glycaemia for the calculation [24]. Artificial intelligence and machine learning were tested in several trials to predict the risk of pancreatic cancer in new-onset diabetes mellitus [30,31].

In addition to new-onset diabetes mellitus (a potential risk factor and/or early sign of pancreatic cancer), other factors influencing the development of the disease must also be considered. They include racial and ethnic differences [32] as well as a combination of other non-modifiable (e.g. age, genetics) and modifiable factors (e.g. lifestyle, smoking, alcohol consumption) [3,33-36].

Attention has been paid not only to new-onset diabetes mellitus but newly also to deteriorating diabetes. Preliminary results from the PANDOME study were published recently [37]. The authors followed 625 individuals over six years and enrolled two cohorts: 97 patients with new-onset diabetes mellitus and 12 subjects with deteriorating diabetes. The second cohort was older, with higher HbA1c concentrations, substantial weight loss, and had insulin requirements. Four biopsies of the pancreas were performed for suspicious lesions, one case of stage-1 PDAC was diagnosed in deteriorating diabetes (0.95; 1/109) [37].

Pancreatic cancer is associated with a requirement for changes of medication one to two years prior to clinical diagnosis [38]. These patients typically start antidiabetic or anticoagulant medications and discontinue antihypertensives. The two-year PDAC risk multivariable-adjusted hazard ratio for starting antidiabetics and cessation of antihypertensives was 4.86 [38].

To highlight the strengths of our analyses, the main advantage of our work is the large long-term and robust cohort of patients diagnosed with new onset diabetes mellitus and prediabetes (treated by metformin). This was made possible by accurate data from national registries. However, we are also aware of certain limitations. The registries do not allow us to identify prediabetes without any pharmacotherapy.

In conclusion, the incidence of pancreatic cancer in the Czech Republic is among the highest in Europe. One-fifth of PDAC cases are diagnosed following new-onset diabetes mellitus in patients over sixty. Unintended significant weight loss combined with new-onset diabetes must not be overlooked, as these can be early signs of PDAC. An individualised diagnostic work-up should follow without any unnecessary delay. Risk scoring models are helpful for a personalised approach.

## Data Availability

Agregated data are available in the supplementary material and on the Zenodo website.

https://doi.org/10.5281/zenodo.21802739

## Author Contributions

Jan Bures and Ondrej Majek conceived and developed this study design, collected, analyzed, and interpreted the data, and wrote and revised the manuscript.

Katerina Hejcmanova and Tereza Dianova provided technical support and retrieved and analyzed data. Ondrej Ngo secured project administration and validation and wrote and revised the manuscript. Darina Kahoutova, Radek Pohnan, Jan Skrha,Stepan Suchanek, Petr Urbanek, Ladislav Dusek, Miroslav Zavoral provided expert support and critical feedback for the revisions of this study. All authors were involved in the critical review of the results and have contributed to, read, and approved the final manuscript.

## Acknowledgements

The work was supported from Programme JAC - project SALVAGE (CZ.02.01.01/00/22_008/0004644) financed by the Ministry of Education, Youth and Sports – Co-funded by the European Union. The authors also thank the RECETOX Research Infrastructure (No LM2023069) financed by the Ministry of Education, Youth and Sports for supportive background.

The authors are grateful to Ian McColl, MD, PhD for assistance with the manuscript.

## Consent

The authors have nothing to report.

## Data Availability Statement

All data generated or analysed during this study are included in this article. More detailed data on the main analysis in this article can be found on the Zenodo platform https://doi.org/10.5281/zenodo.21802739.

---

**Supplementary Table S1. Detailed definition of diabetes mellitus in the NRRHS**

Diabetes mellitus was identified using diagnosis codes (ICD-10), medication codes (ATC), national healthcare procedure codes, and diabetes-related medical device groups. The presence of at least one criterion listed below was sufficient for classification as diabetes.

## S1a. Pharmacological definition (ATC classification)

- A10 – Antidiabetic medication *(with the exception of empagliflozin and dapagliflozin prescribed for chronic heart failure)*

### S1b. Healthcare procedures

- 01201 – Care of a stabilized, compensated type 2 diabetic by a general practitioner
- 01204 – Management of a patient with prediabetes by a general practitioner
- 01299 – Patient referred to a diabetologist for long-term monitoring
- 06129 – Training and instruction on insulin administration
- 06130 – Treatment of hyperkeratosis and pre-ulcerative lesions in diabetics
- 06131 – Special treatment of diabetic ulcerations
- 06145 – Re-education of a patient with diabetes mellitus and their close relative
- 06335 – Training and instruction on insulin administration
- 06637 – Training and instruction on insulin administration
- 13024 – Examination for the risk of diabetic foot syndrome
- 13026 – Evaluation of glycaemic profiles from a glucometer using a computer
- 13051 – Targeted education of a diabetic patient
- 13053 – Team-structured group education for diabetics, for a group of up to 6 people, 180 min.
- 13055 – Treatment of a patient with diabetic foot syndrome (1 foot)
- 13073 – Creation of special contact fixations and splints for diabetic foot syndrome
- 13075 – Professional continuous monitoring using a glucose sensor
- 13077 – Repeated continuous glucose monitoring using a sensor
- 13081 – Optimization of insulin pump settings
- 13082 – Examination of a patient at risk of impaired hypoglycaemia recognition
- 13083 – Setting of a bolus calculator for flexible insulin dosing
- 91801 – Insertion of an insulin pump

### S1c. Diagnoses (ICD-10)

Diabetes mellitus

- E10–E14

Diabetes-related conditions

- G59.0 – Diabetic mononeuropathy
- G63.2 – Diabetic polyneuropathy
- H28.0 – Diabetic cataract
- H36.0 – Diabetic retinopathy
- M14.2 – Diabetic arthropathy
- N08.3 – Glomerular disorders in diabetes
- U69.74 – Diabetic foot syndrome
- Y42.3 – Adverse effects of insulin and oral antidiabetics

### S1d. Medical devices

- Diabetes-related medical devices (Group 05; Group 11 before 2020)

**Supplementary Table S2.** Recent diabetes mellitus among individuals diagnosed with pancreatic cancer in 2022. Diabetes mellitus was defined exclusively based on antidiabetic medication use.

| **Time of diagnosis of diabetes mellitus (DM)** | **39 years and under** | **40-59 years old** | **60-74 years old** | **75 years and over** | **TOTAL** |
| --- | --- | --- | --- | --- | --- |
| DM diagnosed within 1 year prior to cancer diagnosis | 1 (4.8%) | 22 (6.5%) | 85 (7.3%) | 61 (6.0%) | 169 (6.6%) |
| DM diagnosed within 2 years prior to cancer diagnosis | 0 (0.0%) | 9 (2.7%) | 37 (3.2%) | 17 (1.7%) | 63 (2.5%) |
| DM diagnosed within 3 years prior to cancer diagnosis | 1 (4.8%) | 5 (1.5%) | 28 (2.4%) | 16 (1.6%) | 50 (2.0%) |
| DM diagnosed within 4 years prior to cancer diagnosis | 0 (0.0%) | 9 (2.7%) | 31 (2.7%) | 15 (1.5%) | 55 (2.2%) |
| DM diagnosed within 5 years prior to cancer diagnosis | 0 (0.0%) | 2 (0.6%) | 21 (1.8%) | 17 (1.7%) | 40 (1.6%) |
| DM diagnosed 6 or more years prior to cancer diagnosis | 0 (0.0%) | 39 (11.6%) | 287 (24.6%) | 292 (28.6%) | 618 (24.3%) |
| DM diagnosed after the cancer diagnosis | 1 (4.8%) | 15 (4.5%) | 43 (3.7%) | 23 (2.3%) | 82 (3.2%) |
| DM not diagnosed | 18 (85.7%) | 236 (70.0%) | 635 (54.4%) | 581 (56.8%) | 1 470 (57.7%) |
| **TOTAL NUMBER OF PATIENTS DIAGNOSED WITH PANCREATIC CANCER (2022)** | **21 (100.0%)** | **337 (100.0%)** | **1,167 (100.0%)** | **1,022 (100.0%)** | **2,547 (100.0%)** |

